# Evaluating the different control policies for COVID-19 between mainland China and European countries by a mathematical model in the confirmed cases

**DOI:** 10.1101/2020.04.17.20068775

**Authors:** Feng Lin, Yi Huang, Huifang Zhang, Xu He, Yonghua Yin, Jiaxin Liu

## Abstract

This study focuses on evaluating the different policies of controlling the outbreak of COVID-19 in mainland China and in some European countries. The study is based on mathematical model which is a modified susceptible-infected-recovered (SIR) model. The model takes death and recovery into consideration which in convenience is called the susceptible-infected-recovered-death (SIRD) model. The criterion for the recovered patients is assumed by COVID-19 nucleic acid testing negative. The mathematical model is constructed by retrospective study. Determination of the parameters in the model is based on the epidemic bulletin supplied by the Chinese Center for Disease Control and Prevention (CDC) and National Health Commission of the People’s Republic of China (NHC) from Jan 16 2020 to Mar 5 2020. The data cover the date when the epidemic situation is reported and the data showed that the epidemic situation is almost under control in China. The mathematical model mainly simulates the active cases and the deaths during the outbreak of COVID-19. Then apply the mathematical model to simulate the epidemic situations in Italy and Spain, which are suffering the outbreak of COVID-19 in Europe. The determination of the parameters for the 2 European countries is based on the data supplied by Worldometers. By comparing the difference of the parameters based on the same mathematical model, it is possible to evaluate the different policies in different countries. It turns out that the relatively easing control policies might lead to rapid spread of the disease.

## 1 Introduction

The current outbreak of coronavirus disease (COVID-19) was first reported from Wuhan, China, on 31 December 2019 [1]. When the confirmed cases in more than 20 countries worldwide raised concern on an international scale, it is closely monitored by researchers and public alike [2–5]. According to the infection, the data of COVID-19 supplied by the Chinese Center for Disease Control and Prevention (CDC) and National Health Commission of the People’s Republic of China (NHC) in Mainland China, the outbreak of COVID-19 in mainland China is almost under control, while in European countries the epidemic situation is severe as the number of deaths increases rapidly [6].

In January the disease spread very fast in China. According to the information supplied by NHC and the Chinese CDC, the number of confirmed cases was 45 on Jan 16 2020, which increased to more than 80000 on Mar 5 2020. As the increasing rate of the active cases is close to 0 in March, it could be deduced that the epidemic situation is almost under control. However the epidemic situation in European countries was severe in March. As there was a relatively complete process from epidemic outbreak to control in China, it could supply a model by retrospective study. A mathematical model is proposed by modified susceptible-infected-recovered (SIR) model, which for convenience is named as susceptible-infected-recovered-death (SIRD) model. The determination of parameters relies on case number data provided by the Chinese CDC which updates the number of active cases both in China and the global per day [7] and NHC which mainly updates the number of the active cases, the close contacts and the recovered cases in China [8]. Using the mathematical model proposed by the epidemic situation in China, the epidemic situation in European countries could also be estimated in the same way. The determination of the parameters in European countries relies on the data from Worldometer [6]. In this study 2 European countries Italy and Spain where the epidemic situation is quite severe are chosen.

The SIR model is an effective model to predict the growth of the confirmed cases [9]. The parameters in the SIR model are usually constant [2–5]. And the mortality is not taken into consideration. So far, the researchers have already taken banning of traffic into consideration. By retrospective study, it is found that the parameters are actually time-dependent. And the mortality may be an important variable during the outbreak of the disease. Even though the death is much less than the infected population, it needs to be considered. In the traditional SIR model, this kind of information is not taken into consideration. With these modifications, a new model proposed in this study is called the Susceptible-Infected-Recovered-Death (SIRD) model.

The mathematical model in this study only contains 4 kinds of variables: susceptible group *S*(*t*), infected group *I*(*t*) (the confirmed cases), recovered group *R*(*t*) (the nucleic testing negative after infection) and the mortality group *D*(*t*). The transportation control is not involved in this model. The mechanism only reflects the spread of disease in the healthy population. The result in this model could be compared with the transportation control which is the actual result.

## 2 Epidemiological modeling

### 2.1 Epidemiological modeling based on retrospective study of China

Before introducing the mathematical model, the observed results are shown in figure 1. The sub-figures are 1(a)-1(d). The observation of the confirmed cases, the death, the close contacts and the recovered cases lasts for 50 days. It began from Jan 16 2020 to Mar 5 2020. Figure 1(a) and figure 1(b) imply that the growth of the close contacts and the confirmed cases slowed down on the 30th day. And figure 1(d) implies that the growth of the deaths slowed down on the 40th day. According to the data, rapid increase of the confirmed cases implies that the growth of it might be in the exponential form [2].

**Figure 1:**
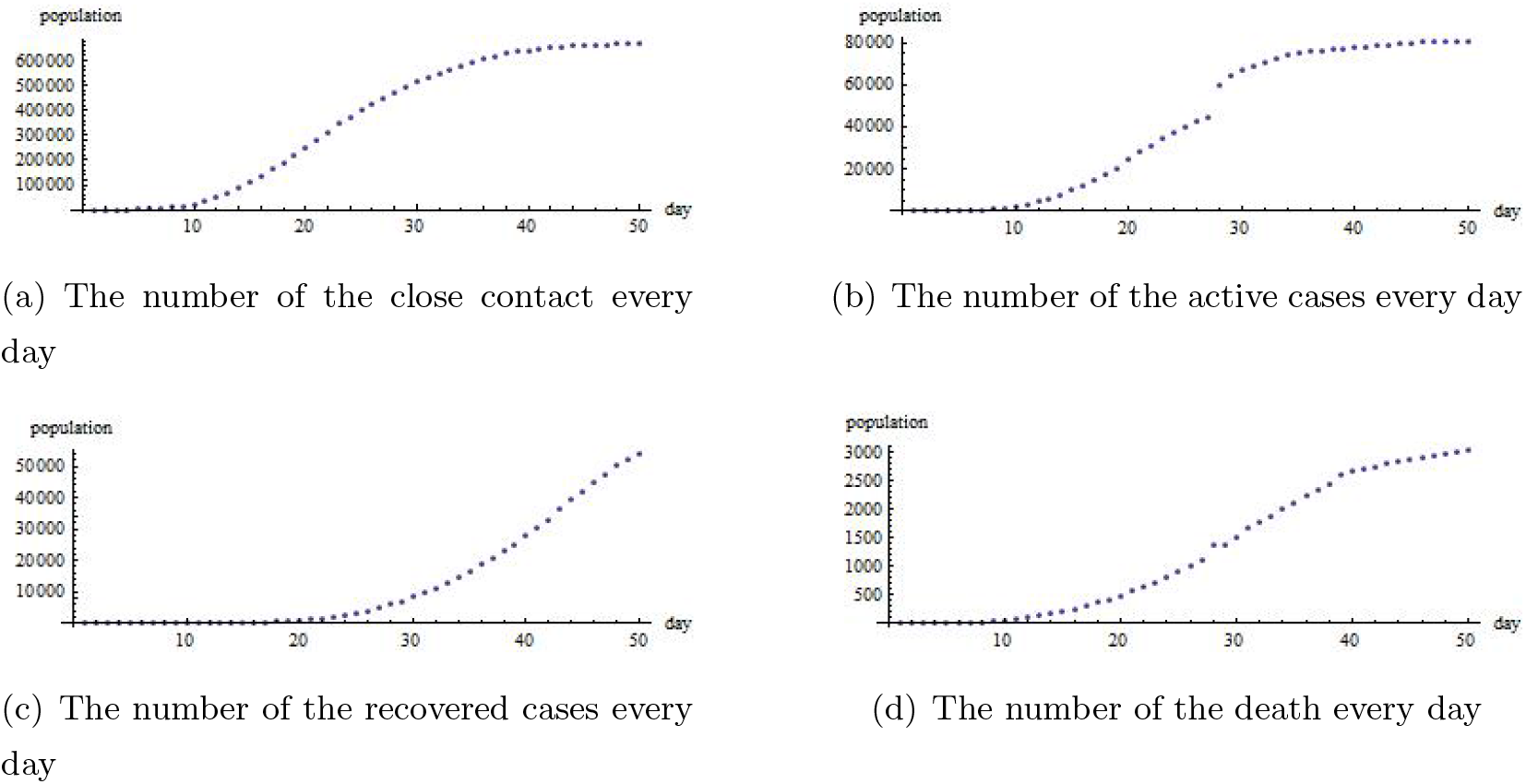
The cumulative number of the close contacts (fig. 1(a)), the active cases (fig. 1(b)), the recovered cases (fig. 1(b)) and the death (fig. 1(d))

Comparing figure 1(a) with figure 1(b), it implies that the close contacts are actually the susceptible population rather than the total population in China or in Hubei province. The transportation policies in China [10–15] is proposed for controlling the outbreak of the disease. If some one was traveling by train as was diagnosed, all the people in that train should be confined in a isolated area. This means that all the people in that train were assumed to be susceptible one, while people who are not in that trip are not regarded as the close contacts. Other policies should also been taken into consideration, such as active cases were either quarantined or put under a form of self-quarantine at home [2]. The suspicious cases were also either quarantined or put under a form of self-quarantine [2]. The policies imply that the number of the susceptible population only depends on the cumulated number of the close contacts.

In this work, the variable *S*(*t*) which denotes the number of the susceptible cases in SIR model is related to the number of the close contacts. By retrospective study, the total number of the close contacts is 6.9*×*10^5^. However, as the epidemic situation is changeable, it is difficult to give the exact number of close contacts, which are the susceptible population. Moreover the number of the close contacts is increasing day by day, while the infected number of population is growing. So it could be deduced that in fact the exact number of the close contacts is unknown. For these reasons, the mechanism of infection-recovering-death is given by (1)-(4):

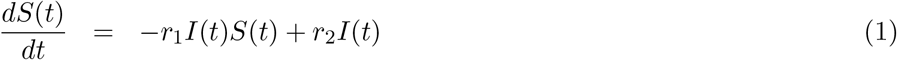

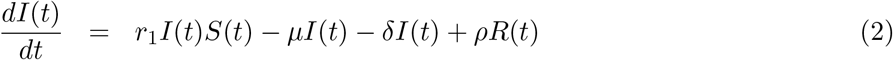

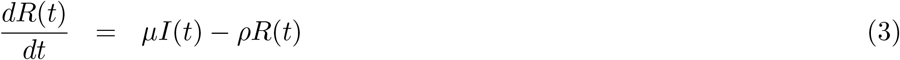

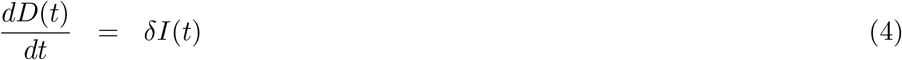

formula (1) means that the number of the susceptible population consists of 2 parts: the number of confirmed cases would increase the number of the susceptible population by the rate of *r*_2_, and the susceptible population is converted into the confirmed cases when they are in contact with the confirmed cases with the rate *r*_1_. When the confirmed cases were in contact with those healthy ones, the healthy ones might be transmitted into the infected cases. The confirmed cases are described as variable *I*(*t*), by formula (2). According to the nucleic testing, the recovered cases could be defined by the infected cases whose nucleic testing negative. In this model, it is assumed that the recovered population with nucleic acid testing positive again might change into the active cases, as it is reported that some recovered COVID-19 patients appeared nucleic acid testing positive [16]. The increase of *I*(*t*) depends on changing the susceptible cases into the active cases and the recovered population with nucleic acid testing positive. The recovery rate *µ* and the death rate *δ* eliminate the number of the active cases. The number of the recovered patients depends on the variable *I*(*t*) in equation (3). The number of the death is described in equation (4). The transmission coefficients would be estimated by the observed results.

### 2.2 Determination of the transmission coefficient

The determination of the transmission coefficients mainly reported in appendix. In this section we only give an introduction of determining the transmission rate. The transmission rate *r*_1_ calculated by the observed results is shown in figure 2(a), and the death transmission rate *δ* calculated by observed results in figure 2(b).

**Figure 2:**
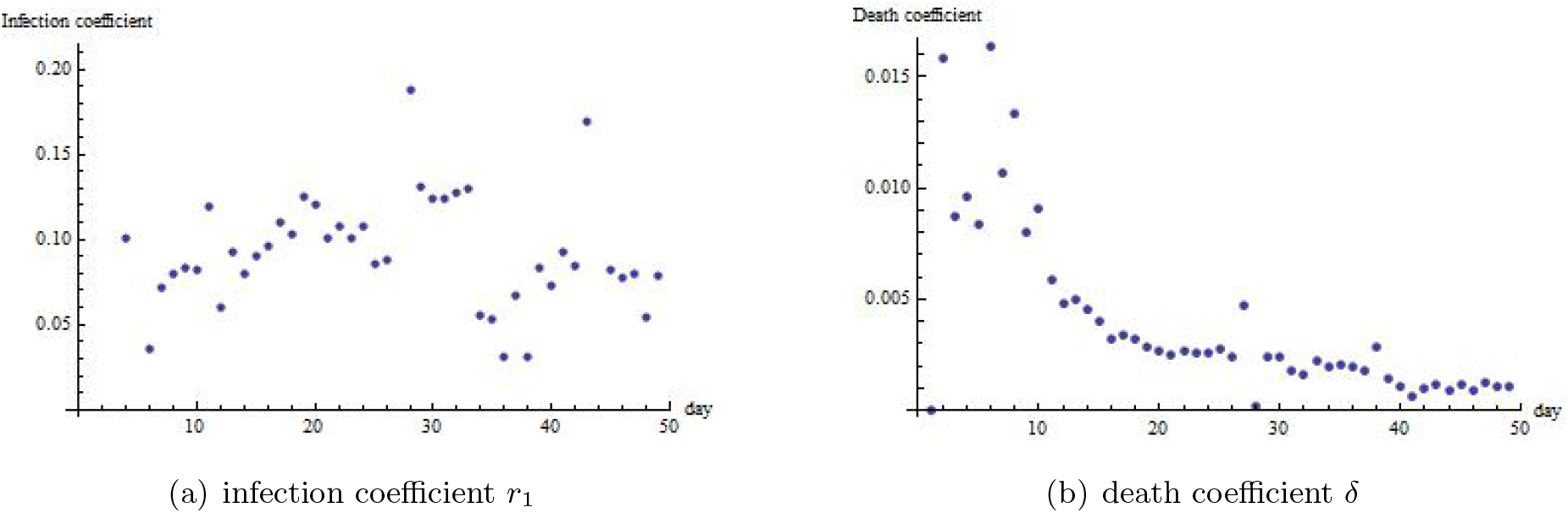
The infection coefficient is figured out in this way: divide the number of the diagnosed people by the number of the close contacts every day. The death coefficient is figured out in this way: divide the number of the death by the number of the recovered patients every day

According to the statistical character of *r*_1_, its mean value is 0.143. The shape of its distribution function skew-right (skewness*>* 0), and the distribution function is “thinner” than normal distribution (kurtosis*>* 3). So the most probable number of *r*_1_ is larger than 0.143. In this work we choose *r*_1_ = 0.145. The detailed approach of determining the formula of the transmission rates would be shown in appendix. Similarly, the recovery parameter is also time-dependent. The details of determination of the time-dependent function of *δ*(*t*) and *µ*(*t*) are shown in appendix. There only show the results. *δ*(*t*) is chosen in the formula as (5).

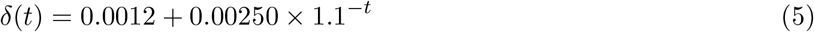

*µ*(*t*) is given in the formula (6)

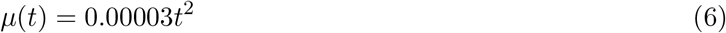

The parameter *ρ* = 0.005, as the incidence of the recovered patients with nucleic acid testing positive is very low. *r*_2_ is to be set according to the observed number. *r*_2_ must let the peak number of the confirmed cases similar to the observed ones.

### 2.3 Prediction of epidemic situation in European countries

As the epidemic situations in European countries is changeable, the model (1)-(4) is also used for prediction. The curing rate is assumed to be the same, as there is no reason to assume the medical resources in European countries are better or worse than it in China. The difference stems from the medical resources management, which are concerned with *δ, r*_1_ and *r*_2_. The data have been in collection since Feb 15 2020 [6]. According to the observed data, the parameters *r*_1_ and *r*_2_ should be adjusted. The death rate needs to be re-estimated as it reflects the management of medical resources in European countries.

Lack of medical resources is also reflected in European countries. Comparing with the cases in China, it might be more severe in European countries. The death rate of Italy and Spain is estimated as in the formula (7):

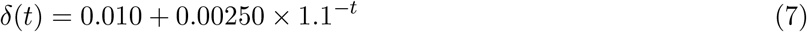

the formula (7) is estimated in the same way as that was done for China. It is obvious that the death rate is larger in European countries than that in China.

## 3 Results

### 3.1 Death rate in China, in Hubei province China, in Italy and in Spain

Hubei province contains the largest number of the infected population in China. And most of the confirmed cases are in Hubei province. This is the result of the policies that confining the migration of the people during the outbreak of the COVID-19. The tendency of the death in Hubei province is consistent with the death in mainland China (figure 3)

**Figure 3:**
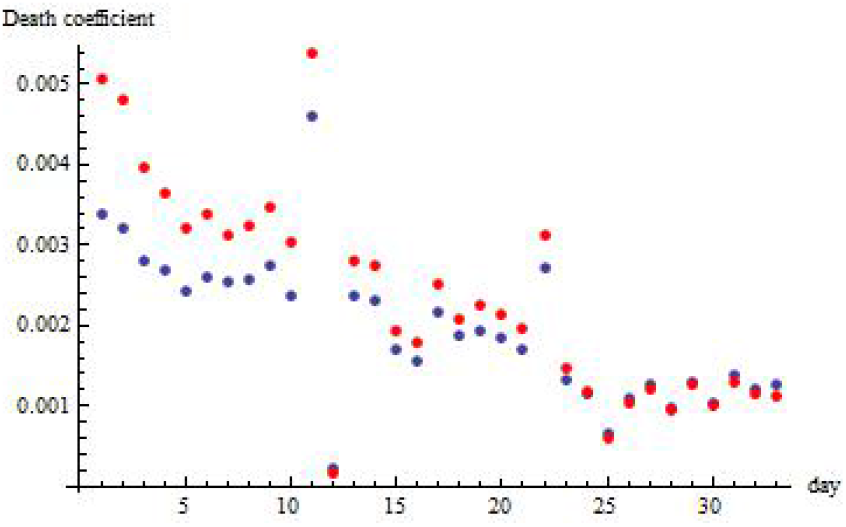
The red points are the death coefficients in Hubei province and the dark blue points are the death coefficients in China

Figure 3 implies that the evolution of the SIRD model might show the same results in Hubei province. And it could be deduced that if the epidemic situation shows the outbreak, the tendency of the death coefficients are the same in both China and Hubei province China. As lack of experience and medical resources at the beginning, he death coefficient might be high. Then the death coefficient tends to a stable constant.

Lack of medical resources is also reflected in European countries. Comparing with the case in China, it might be more severe in European countries. The death rate of Italy and Spain is estimated as in the formula (7). It is obvious that the death rate is larger in European countries than in China. The fitting line is shown in figures 4(a)-4(b)

**Figure 4:**
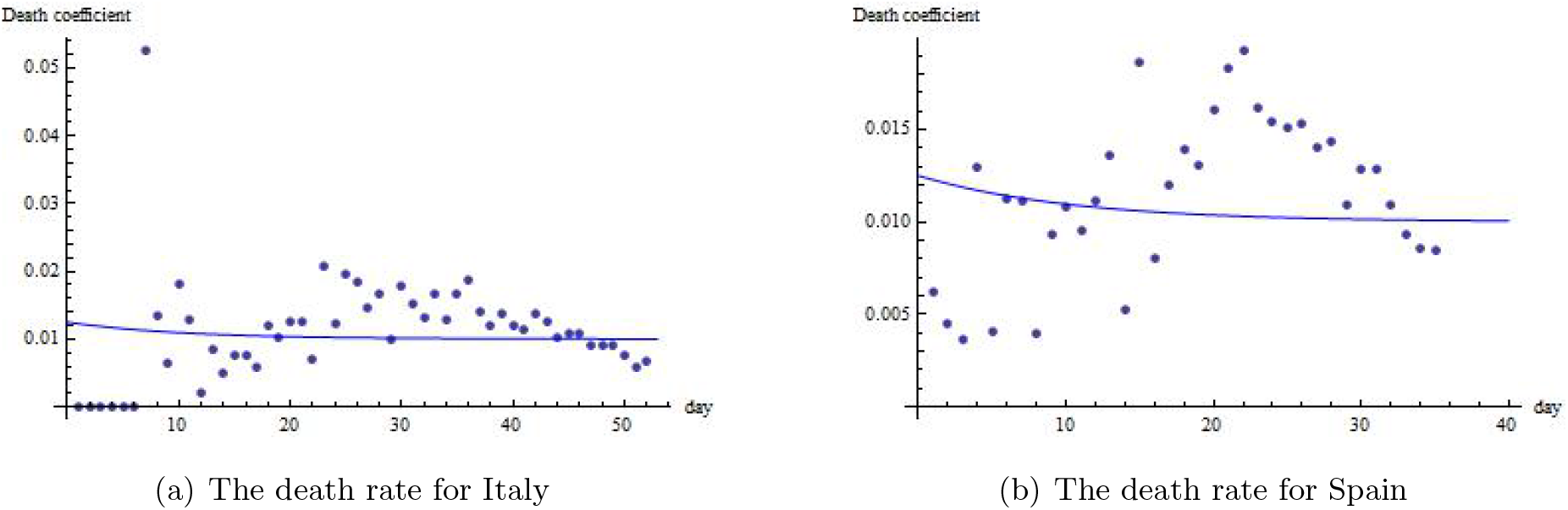
The death rate figured from the observed data was in dots, and the simulated death rate is drawn in blue line. As at the beginning of the 18 days, there were no reported deaths in Spain, the estimation of Spain began from 3th Mar 2020

### 3.2 The simulated result of China

At initial time *t*_0_, the number of the recovered patients is *R*(*t*_0_) = 2, the number of active cases is *I*(*t*_0_) = 45. The number of the susceptible population is *S*(*t*_0_) = 763. By numerical solving equations (1)-(4), the simulation of the number of *S*(*t*), *I*(*t*), *R*(*t*) and *D*(*t*) is shown in figure5. *S*(*t*) is the black dashed line in figure 5(a). The line is not clear in figure 5(a). As the number of the susceptible population is changeable because of the number of the active cases, and it is not important in this study, it is not shown independently in figure 5. *R*(*t*) is the brown dashed line in figure 5(a). The reason why it is much larger than the active cases is that the number might be recounted. To show *I*(*t*) and *D*(*t*) clearly, *I*(*t*) is shown in figure 5(b) and *D*(*t*) is shown in figure 5(c):

**Figure 5:**
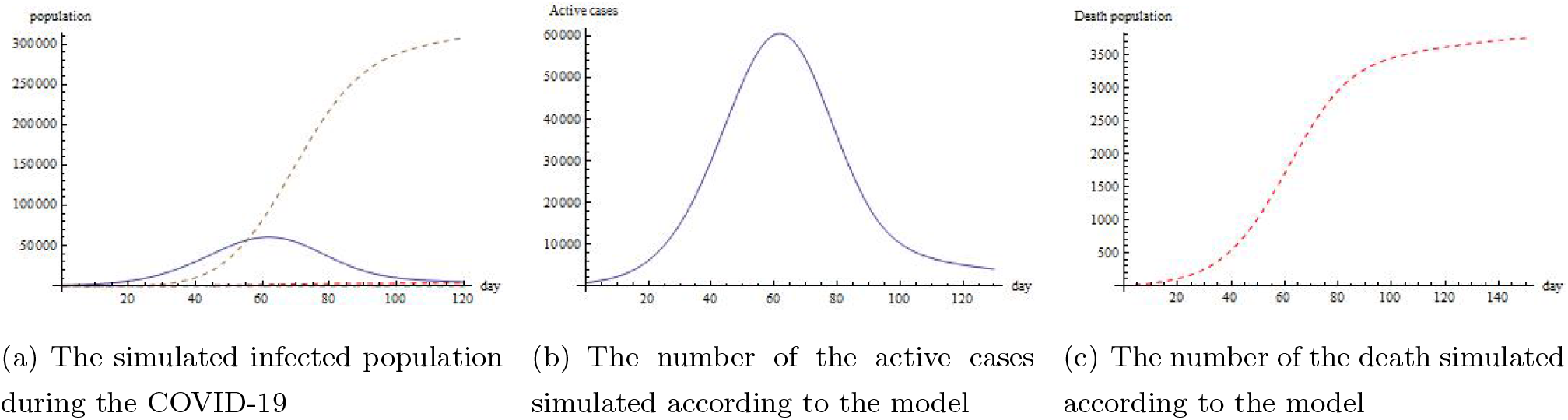
The simulated COVID-19 infected population in China. The blue line (fig. 5(a) and fig.5(b)) is the number of the population that infected by the disease. The brown line (fig. 5(a)) is the population that are recovered. And the dashed red line is the number of the death (fig. 5(a) and fig.5(c)). The parameters are *r*_1_ = 0.145, *r*_2_ = 0.107, *ρ* = 0.005. *µ*(*t*) and *δ*(*t*) are referred to formular (6) and formula (5). The initial conditions are *S*(*t*_0_) = 763, *I*(*t*_0_) = 45, *R*(*t*_0_) = 2, *D*(*t*_0_) = 1

#### 3.2.1 Comparing with the actual results

Comparing the simulated results with the actual results would reflect the infection under 2 kinds of cases: one is the ad libitum transportation, the other is the transportation under control. It has already been mentioned in the previous introduction, the mechanism of this model did not take the transportation control into consideration. So this result could reflect how the infection would evolve without transportation control. Figure 6 shows the comparison of the simulated number with the actual number supplied by the Chinese CDC. Figure 6(a) gives the comparison of the number of the infected population every day in mainland China with the simulated result. And figure 6(b) shows the comparison of the number of the death in mainland China (blue dots), Hubei province China (red dots) and the simulated result. In fig.6(a), the data of the active cases in Hubei province China is limited, so the number of active cases is not available.

**Figure 6:**
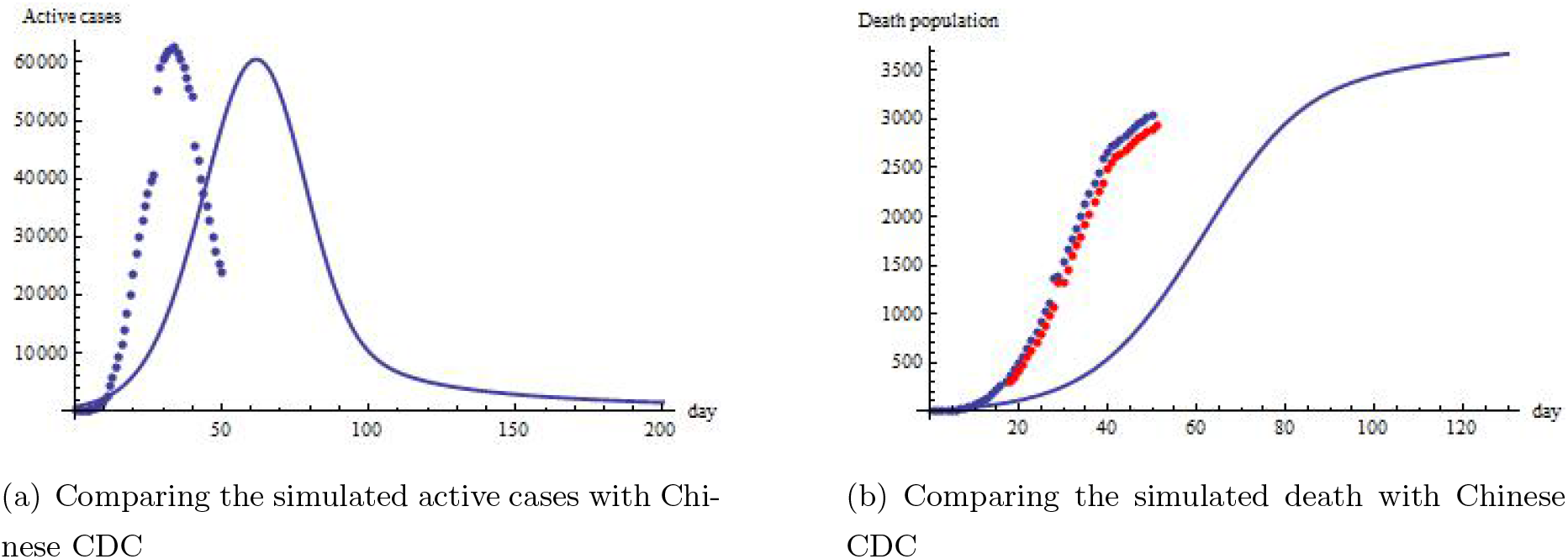
The blue dots denotes the number of the active cases (fig.6(a))/ death (fig.6(b)) by COVID-19 in China every day. The red dots denotes the number of the death (fig.6(b)) by COVID-19 in Hubei province China every day. The number of the active cases in Hubei province China is not available.

Generally, the increase of the active cases and the death simulated from the mathematical model is not as large as that in the actual data, and the key performances such as the peak number of the active cases, the number of the death are consistent with the actual information.

### 3.3 Simulation result of Italy and Spain

In European countries, the mechanism is the same as that in China. The time-dependent parameter the recovery rate is the same as that in China. The difference stems from the parameters like *r*_2_, *ρ* in the model. The death rate might be high at the beginning, because of lack of medical resources and curing experience. Later the death rate is slowed down, while the recovery rate would increase slowly. The policies in European country is different from that in China. The prediction results of Italy are shown in figure 7(a)-7(b).

**Figure 7:**
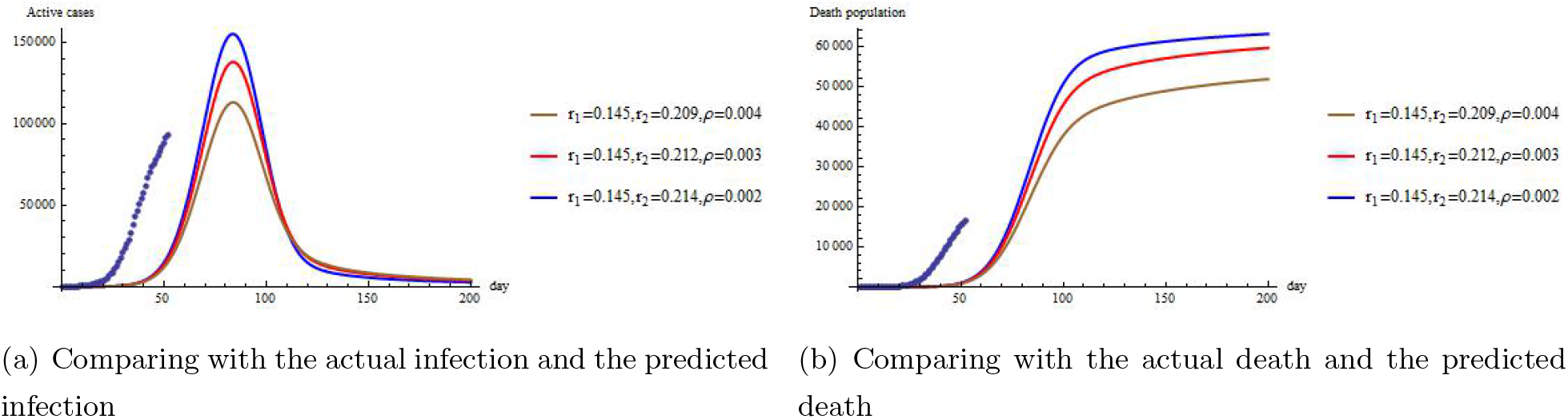
The predicted COVID-19 infected population in Italy. The dots in figures 7(a)-7(b) are the actual results. Figure 7(a) is the simulated number of the active cases. Figure 7(b) is the simulated number of the death. The parameters that are not shown in figures 7(a)-7(b) are *µ*(*t*) and *δ*(*t*) which could be referred to formula (6) and (5). The initial conditions are *S*(*t*_0_) = 0, *I*(*t*_0_) = 3, *R*(*t*_0_) = 0, *D*(*t*_0_) = 0

Similarly, the prediction result simulated for Spain is shown in figure 8(a)-8(b). The simulation for Spain began on Feb 24 2020 when the first case was confirmed.

**Figure 8:**
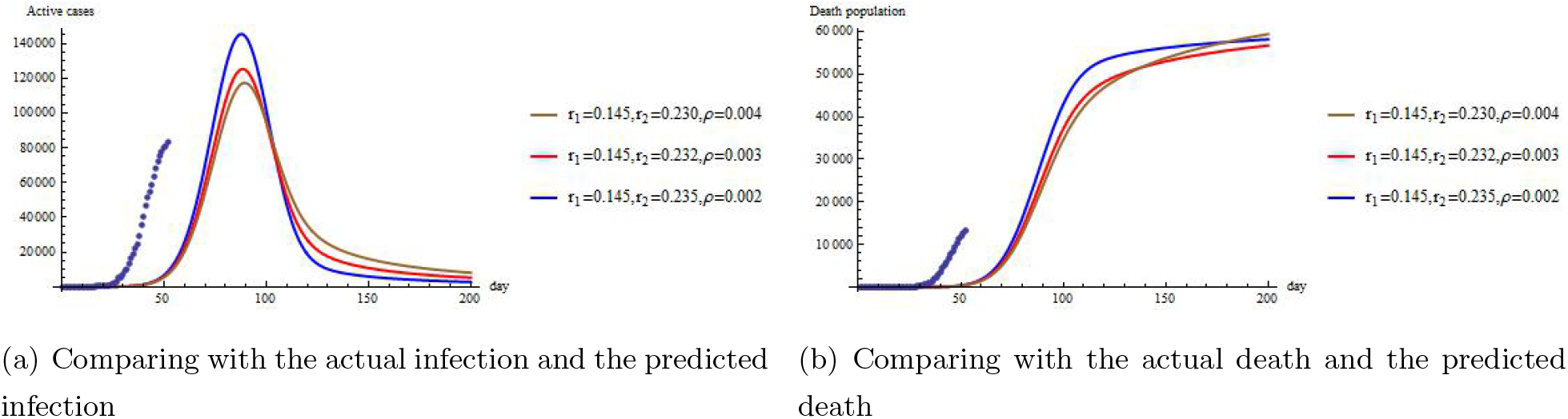
The predicted COVID-19 infected population in Italy. The dots in figures 8(a)-8(b) are the actual results. Figure 8(a) is the simulated number of the active cases. Figure 8(b) is the simulated number of the death. The parameters that are not shown in figures 8(a)-8(b) are *µ*(*t*) and *δ*(*t*) which could be referred to formula (6) and (5). The initial conditions are *S*(*t*_0_) = 0, *I*(*t*_0_) = 1, *R*(*t*_0_) = 0, *D*(*t*_0_) = 0

Similar to the simulation in China, the increase rate in simulation is not as large as that in the actual data. As it has been reported that the susceptible cases might be very inexplicit [17–19], the initial number of the susceptible cases *S*(*t*_0_) is not accurate. It would be shown in the section of discussion.

## 4 Discussion

### 4.1 Comparing the control policies between China and European countries

The model simulation for China and European countries are with the same mechanism. The only difference are the parameters *r*_2_ and *ρ*. And it results in the difference of *δ*. According to equations (1)-(4), *r*_2_ is related to the increasing of the susceptible cases and *ρ* is related to the number of the recovered patients with nucleic acid testing positive. Both of the parameters are related to the transportation policies. The parameters are determined by the actual data, so the simulation in figures 6-8 are close to the actual data. The parameter *r*_2_ and *ρ* are adjusted to fit the observed data. Though the simulation result is different from the observed data, mainly in the increase rate, the relative quantity of *r*_2_ and *ρ* in China and European countries implies whether the epidemic control policies is rigorous.

### 4.2 The initial number of the susceptible cases might not be accurately determined

In figures 6-8, it is clear that the simulation result is delayed comparing with the actual result. There might be 2 reasons: one is that the mechanism does not explicitly contain banning of the transportation [12, 15, 20], the other is that the initial number of susceptible cases is not accurate due to lack of access to COVID-19 testing in the early weeks [21–23]. For the second reason, we change the initial number of *S*(*t*_0_) to check whether this deduction is right.

If in fact the initial number of the susceptible cases was 3000 rather than 763 in China, and was 300 rather than 0 in Italy and Spain, then the active cases and the death is shown in figures 9-10.

**Figure 9:**
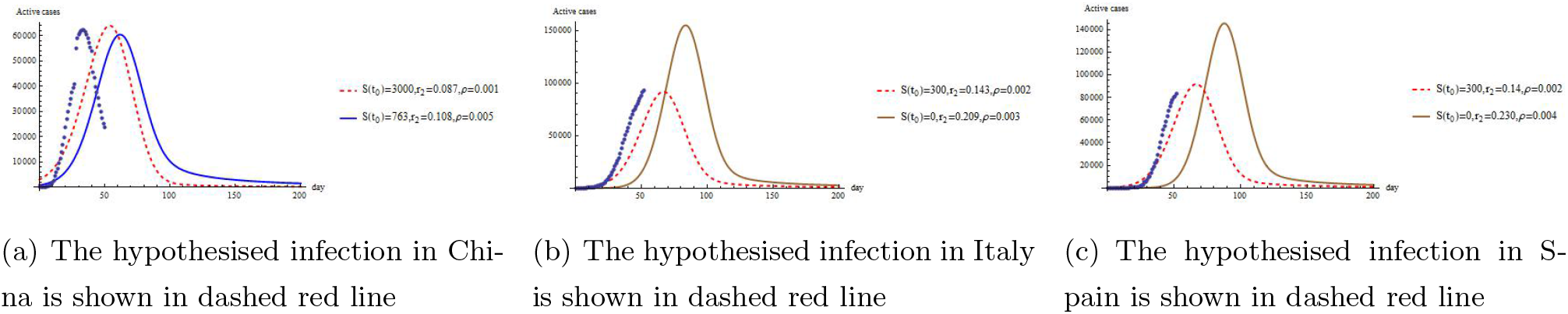
All the parameters and initial conditions except those denoted in figures 9(a)-9(c) remain the same as those in the previous simulation of number of the active cases. The dashed red lines show the simulation with hypothesised *S*(*t*_0_).

**Figure 10:**
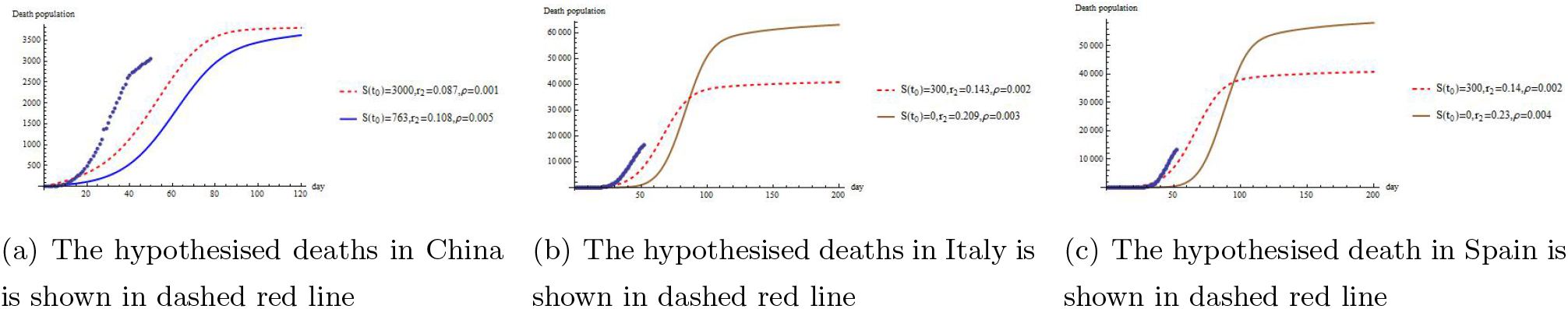
All the parameters and initial conditions except those denoted in figures 10(a)-10(c) remain the same as those in the previous simulation of number of the death. The dashed red lines show the simulation with hypothesised *S*(*t*_0_).

The number of the active cases were the same as those in the previous simulation, the only change is the initial number of the susceptible population and the coefficient *r*_2_. It turns out that *r*_2_ for European countries is still larger than that for China. It implies that the banning of transportation for China is more rigorous. Moreover, the initial number of the susceptible population is larger than that was reported regardless of the country due to lack of access to COVID-19 test kits in the early weeks.

In view of the simulation by mathematical model, the initial number of the susceptible cases might be difficult to be determined. This might be demonstrated by the report that 60% of active cases might have no symptoms but could infect other healthy people [17–19]. And it is reported that the infected cases are of strong infection when they appeared to have no symptoms [24]. So when the first infected person appears in some area, the infected ones with no symptoms might convert the healthy people into the susceptible ones without records. In the hypothesised simulation, the actual data seemed to be closer to the simulated curves. But the increasing rate of the simulation is not as large as that in the observed data. The reason might be that the transportation control was not considered in this mechanism, which has already been mentioned in the previous section. This implies that banning of the traffic is an effective way to prevent the spreading of disease.

## Data Availability

The original data available could be referred to the links:
http://2019ncov.chinacdc.cn/2019-nCoV/
http://2019ncov.chinacdc.cn/2019-nCoV/
https://www.worldometers.info/coronavirus/

http://www.nhc.gov.cn/xcs/yqtb/list_gzbd.shtml

https://www.worldometers.info/coronavirus/

## Author contributions

Feng Lin performed the research. Feng Lin, Jiaxin Liu and Yi Huang designed the research study. Feng Lin, Xu He and Yi Huang analyzed the data. Yonghua Yin, Feng Lin, Yi Huang and Huifang Zhang wrote the paper.

## 5 Appendix: determination of the coefficient

### 5.1 The infection coefficient in China

The infection coefficient is calculated in this way:

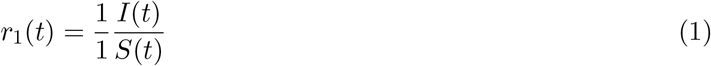

The reason why we wrote 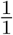 before the transition rate of infection is that it is assumed that the contact in fact occurs one by one quite often. The dimension of 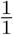 is per person i.e.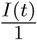 is dimensionless. For calculation convenience, we just figure out the variable part. According to the data given by Chinese CDC and NHC, the bound of infection coefficient is *r*_1_ ∈ (0.02, 0.21). It is hard to give a formula to describe *r*_1_. The best way is to choose an exact number in its 95% confidence interval to set *r*_1_. The statistical character of infection coefficient is given by table 1. The confidence interval is figured by the population mean estimated from the actual data:

**Table 1:** The statistical character of the infection coefficient

| Table 1: The statistical character of the infection coefficient |  |  |  |  |
| --- | --- | --- | --- | --- |
| mean | variance | skewness | kurtosis | 95% C.I. |
| 0.143738 | 0.0483431 | 4.82231 | 27.0367 | (0.0791815, 0.208294) |

### 5.2 The death coefficient in China

The death coefficient is calculated in this way:

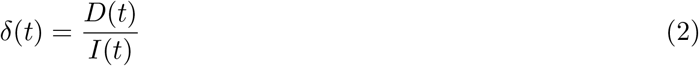

The death coefficient is like some exponential function of *t*. By fitting the time to the ln *δ*(*t*). It changes from time to time. However, the tendency of the death coefficient is like some exponential function of *t*. So by changing the bases of exponential function, it could turn out a series of fitted function. Some fitted function is shown in figure 11

**Figure 11:**
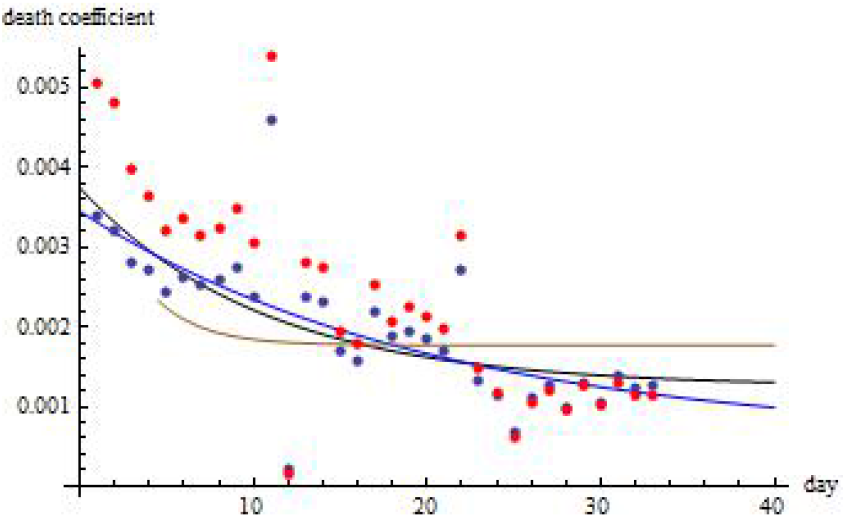
The fitted lines correspond to different base of exponential function. The blue dots are the death coefficient in the mainland of China, and the red dots are the death coefficient in Hubei province China. The data of the death and the patients could be checked on the website of Chinese CDC and NHC.

**Figure 12:**
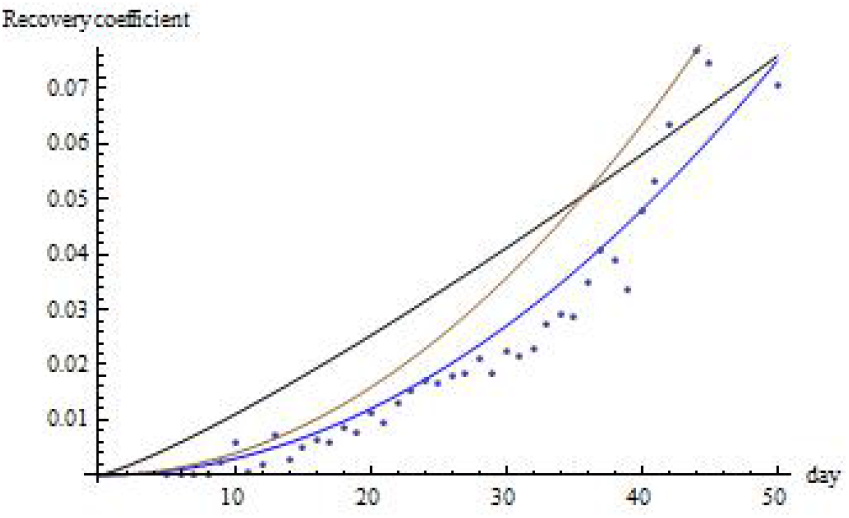
The recovery coefficient per day is shown in dots, and the simulated curing coefficients is shown in the line

In figure 11, the black line is:

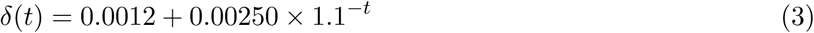

the brown line is

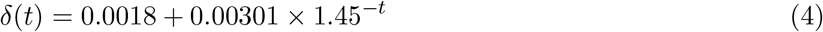

The fitted formulas are given below:

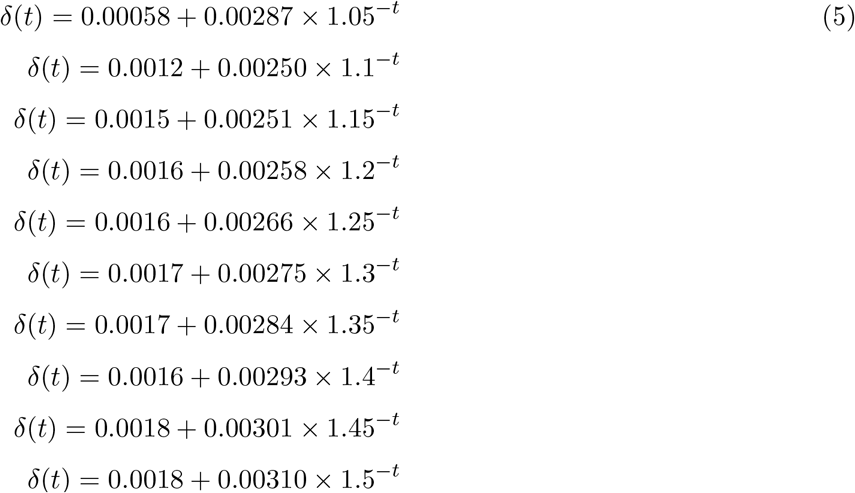

In this study we choose the formula fitted as black line:

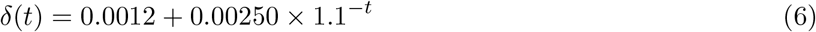

the reason is that when *t* goes to infinity, *δ →* 0.0012.

### 5.3 The recovery coefficient in China

The recovery coefficient is calculated in this way:

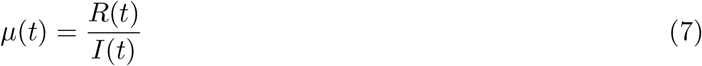

*µ*(*t*) is like some power function of *t*. By changing the power of *t*, the result gives:

The fitted formula of black line is

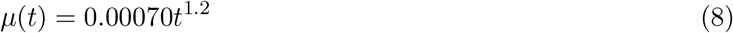

The brown line is the fitted formula

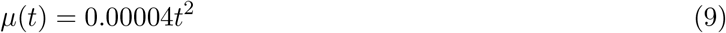

however, the blue line is the best:

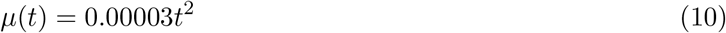

The fitted formulas are given below:

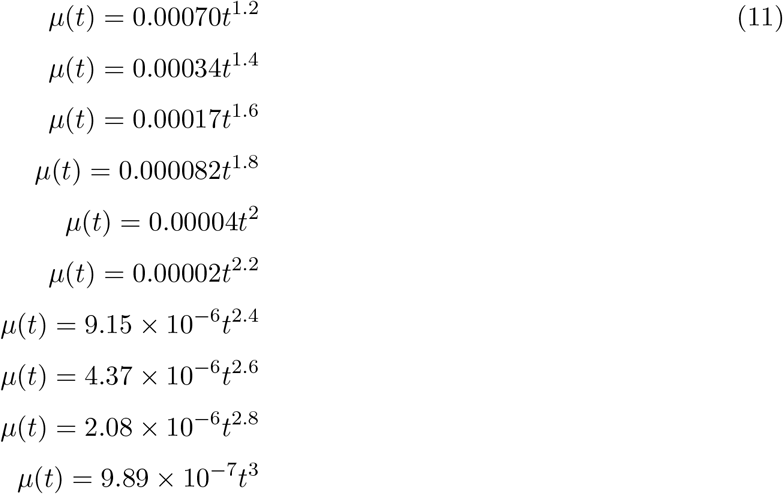

In this study we use the formula:

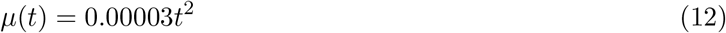

